# Assessment of pain types in recently diagnosed patients with inflammatory arthritis

**DOI:** 10.1101/2025.04.15.25325860

**Authors:** Z. Rutter-Locher, S. Norton, J. Taylor, B. Menon, T. Esterine, R. Williams, L.S. Taams, K. Bannister, B.W. Kirkham

## Abstract

**Objective:** Up to 40% of patients with inflammatory arthritis (IA) experience persistent pain, traditionally thought to be associated with a shift from peripheral to centrally mediated pain during the disease course in some patients. We assessed sensory profiles of recently diagnosed individuals with IA, hypothesising that pain reported at this early stage of diagnosis is driven predominantly by peripheral joint inflammation.

**Methods:** Recently diagnosed IA patients with pain numerical rating scale (NRS) scores of ≥3 were recruited. We collected data on: Arthritis activity (Disease Activity Score-28 (DAS-28), musculoskeletal ultrasound); quality of life (Musculoskeletal Health questionnaire (MSK-HQ), Euro Quality of Life questionnaire (EQ-5D)); mental health status (Patient Health Questionnaire Anxiety, Depression Scale (PHQ-ADS)), and pain characteristics (fibromyalgia criteria, painDETECT, Static and Dynamic Quantitative Sensory Testing, QST).

**Results:** 61 participants (57% female, 62% rheumatoid arthritis) were enrolled: mean age 49.8 ±15; time since diagnosis 1.2±2.3 months. 97% had peripheral joint inflammation, with a mean DAS-28 score of 3.8±1. However, 20% had a tender minus swollen joint count of ≥7, 21% met the fibromyalgia criteria and 25% had a painDETECT score of ≥19, which significantly correlated with DAS28, MSK-HQ and PHQ-ADS scores. QST revealed lowered pressure pain thresholds at non-articular sites in a subset of participants and facilitated temporal pain summation and deficient pain modulation in 18% and 61% of patients respectively.

**Conclusion:** This study provides evidence of centrally mediated pain at the time of diagnosis, challenging the notion that, even at the early stage of disease, pain is driven only by peripheral mechanisms.

**SIGNIFICANCE AND INNOVATIONS:**

- This study is the first to record comprehensive sensory profiles incorporating clinical examination, ultrasound, questionnaire, and quantitative sensory testing data in the early stage of inflammatory arthritis diagnosis.
- Of those tested, 97% had peripheral joint inflammation confirmed by clinical assessment and ultrasound. In addition, clinical, patient-reported and QST outcome measures revealed the likely presence of centrally mediated pain in a subset of participants.
- These findings challenge the current belief that only peripheral mechanisms driven by joint inflammation underpin pain in the early stage of disease progression, highlighting a role for centrally mediated pain even in the early stage of disease.

## INTRODUCTION

Despite major improvements in therapies and treatment strategies, up to 40% of patients diagnosed with inflammatory arthritis (IA), including rheumatoid arthritis (RA), psoriatic arthritis, and ankylosing spondylitis, continue to experience persistent pain (1–4). Pain intensity initially improves during the six-month period post-diagnosis, likely attributable to the anti-inflammatory properties of disease-modifying anti-rheumatic drugs. However, after that period, joint tenderness and pain persist, which might be unresolved inflammation, or as often found in the absence of synovitis it might be pain sensitisation (1), or a combination of both (5). A recent meta-analysis by our group found that, according to patient-reported outcome measures, pain sensitisation was present in approximately 30% of individuals with established IA, with the largest group being those with psoriatic arthritis (9). What underlies this pain sensitisation remains equivocal, with postulated mechanisms encompassing dysfunction in both peripheral and central pain processing pathways including spinal and supraspinal circuits (6–9). Improving our understanding how persistent pain is initiated, develops, and is maintained in the IA patient population is crucial since it could help guide appropriate therapeutic strategies.

Pain in IA can have many different causes, ranging from persistent joint inflammation to dysfunctional neural processes, which requires the use of multiple assessment tools to provide a more complete understanding. Musculoskeletal ultrasound (MSK US) provides an objective assessment of peripheral joint inflammation in addition to clinical examination, while quantitative sensory testing (QST) allows activity in peripheral nerves to be inferred (static QST) alongside the functionality of central pain processing pathways (dynamic QST). When combined with validated questionnaire data, which provides insights into the patient pain experience, clinical examination, MSK US and QST outcomes can be used to build an individual’s sensory profile. Cumulatively, these profiles can help deduce likely pain-causing mechanisms (10,11)

Studies using questionnaire and QST methods, mainly cross-sectional design in patients with longstanding IA (12,13), have suggested the presence of centrally mediated pain in a subset of patients. This research has led to the current consensus, that, in some patients, during their disease course, pain primarily caused by peripheral joint inflammation at diagnosis transforms to include pain that is underpinned by dysfunction in central nervous system processes (6). The temporal nature of this shift during disease course remains unclear as no studies have explored detailed pain characteristics at diagnosis, and there are limited longitudinal studies. In keeping with the current understanding that centrally mediated pain develops over the course of the disease, we hypothesised that peripherally mediated inflammatory pain would predominate during this initial disease phase, with little evidence of dysfunctional central pain processing. Here we report the initial sensory profiles of individuals with IA promptly after diagnosis, established with an array of techniques encompassing both MSK US and QST, alongside validated questionnaires. To our knowledge, this is the first study to employ such a detailed, multimodal approach to sensory profiling in newly diagnosed IA patients.

## PATIENT AND METHODS

### Study population

This study presents baseline data obtained from an initial cohort of 61 patients enrolled in the “Pain phenotypes and their Underlying Mechanisms in Inflammatory Arthritis” study (PUMIA). PUMIA is a single site prospective longitudinal observational study aiming to understand pain mechanisms in IA. Recruitment of participants took place at the rheumatology outpatient department of Guy’s Hospital, London, between February 2022 and November 2022. Inclusion criteria for the study encompassed individuals who reported a total numeric rating scale (NRS) pain score of 3 or higher on a scale of 0 to 10, received a clinician diagnosis of peripheral inflammatory joint disease, and experienced symptom onset within the past 12 months. Patients were enrolled in the study as soon as feasible following their diagnosis.

Patients were excluded if they were: under 18 years of age, unable or unwilling to provide informed written consent, unable to adhere to study protocols, pregnant or breastfeeding, receiving non-inflammatory arthritis-related immunosuppressant therapy, undergoing current or recent (last 90 days) treatment with investigational agents, or having a history of severe peripheral vascular disease or peripheral neuropathy. Use of opioids, gabapentin, or pregabalin were stopped 24 hours prior to assessment. All participants gave full informed written consent and the study protocol received approval from Bromley Research Ethics Committee and the Health Research Authority (REC 21/LO/0712).

### Clinical assessment and US

Routine demographic data (including age, gender, BMI, smoking status, employment status and co-morbidities), disease data (including diagnosis, disease duration, serology) and medication history was obtained from each participant. The term “current steroids” refers to oral prednisolone taken within the past two weeks or intramuscular methylprednisolone acetate administered within the last three months.

Patients were assessed for disease activity (CRP, 68Tender joint count, 66Swollen joint count, DAS28), and completed questionnaires for, quality of life (Musculoskeletal Health Questionnaire, EuroQol-5D (14)), impact of disease (Rheumatoid Arthritis Impact of Disease Questionnaire, RAID (15)), somatic symptoms (Patient Health Questionnaire-15 (16)) and mental health (Patient Health Questionnaire-9, PHQ-9 for depression (17), Generalised Anxiety Disorder assessment-7, GAD-7 (18)). Fibromyalgia status was determined according to the 2016 revision of the ACR 2010 modified fibromyalgia diagnostic criteria (19) (Widespread Pain Index (WPI) and Symptom Severity Scale score (SSS)). Fibromyalgia severity was assessed as the sum of the WPI and SSS scores. Pain assessment was conducted using 1) the NRS for pain where individuals are asked to score, between 0-10, both total body pain and most painful joint pain in the last 24 hours (the ‘target joint’) and 2) the painDETECT questionnaire, which identifies neuropathic-like pain (20). The painDETECT has been used in previous studies as a proxy for pain caused by central mechanisms due to the similarity in symptom profiles (21–23).

A 14 joint MSK US assessment was used to obtain an objective evaluation of joint inflammation, particularly subclinical synovitis. Bilateral radiocarpal, intercarpal, ulnocarpal, MCP 2 and 3, PIP 2 and 3 and MTP 2 and 3 joints were graded semi-quantitatively for grey-scale (GS) and power doppler (PD) according to the EULAR-OMERACT criteria (24) by two individual raters, blinded to patient demographics or sensory profile.

### Quantitative sensory testing (QST)

#### i. Static QST

The QST protocols established by the German Research Network on Neuropathic Pain (DFNS) were applied to assess cutaneous sensory detection using standardised von Frey hairs (Mechanical Detection Threshold, MDT) and pain thresholds using a standardized pinprick set (Mechanical Pain Threshold, MPT) on the target most painful joint and control site (contralateral volar forearm) (25). These tests are performed using standardised instructions where normative data, grouped by age, gender and site is available to allow comparisons with healthy controls (26). The established DFNS protocol was also used to assess pressure pain threshold (PPT) at the joint line of the target most painful joint, the dorsal aspect of bilateral wrists and a non-articular control site (bilateral trapezius muscles) using a Wagner Force 25 FDX algometer (25). PPT is the point at which a pressure applied first becomes painful and lower pain thresholds reflect higher pain sensitivity. Lower PPT at non-articular sites suggest widespread pain likely underpinned by central mechanisms while lower PPT at joint sites only suggest local joint pain sensitivity due to joint inflammation/peripherally mediated mechanisms.

#### ii. Dynamic QST

Dynamic QST comprises Temporal summation of pain (TSP) and Conditioned pain modulation (CPM) paradigms. The TSP paradigm provides a proxy measure of spinal neuronal activity. In brief, delivery of an equal-intensity noxious stimulus above a critical rate leads to an increase in the individual’s perception of that stimulus. We used two (TSP) paradigms. The first followed a DFNS protocol using pinprick stimulators (25) and the second followed a standard protocol using cuff pressure algometry (Detailed protocol in Supplementary material). Spinal neuronal activity can be modulated through descending modulatory circuits, whose function can be assessed upon application of a CPM paradigm (12). In brief, a measure of painfulness for a test stimulus is taken, in the absence or presence of a second painful ‘conditioning’ stimulus. The difference in the participant’s rating of the test stimulus (before and during conditioning) is the measure of a ‘CPM’ effect, where a ‘CPM responder’ refers to an individual who experiences a significant reduction in pain rating of the test stimulus upon conditioning while a ‘CPM non responder’ does not. For the CPM paradigm, we followed a computerised cuff algometry protocol (27)

### Statistical analysis

All data are presented as mean ± standard deviation (SD). Paired or independent t-tests were employed, as appropriate, to identify significant differences in means, with p-values below 0.05 considered statistically significant. To normalize the data for MDT, MPT and TSP, logarithmic transformations were applied, resulting in values with a mean of 0 and SD of 1 (26,28). Absolute reference data (matched for age, gender and site, being the dorsum of the hand) was used to normalise test results of the individual participant by calculating the z transformation= (value participant)- mean controls)/SD controls (29). A z-score of ±1.96 corresponded to statistical significance at the 0.05% level.

Pearson correlations were used to investigate the relationships between markers of central pain mechanisms, peripheral inflammation, and QST parameters with clinical outcomes, such as pain levels, disease activity, and mental health. Given the exploratory nature of this analysis, effect sizes were primarily evaluated, and p-values below 0.05 were considered statistically significant.

Finally, an exploratory factor analysis (EFA) was conducted to identify underlying latent variables (factors) accounting for the common variance across observed variables (Detailed protocol in supplementary material) All analyses were performed using STATA 17.0.

## RESULTS

### Demographics and clinical assessment

#### Demographics

We recruited 61 patients comprised of a broad range of ethnicities, predominantly with a diagnosis of RA (62.3%), with the remaining 38% a mixture of early inflammatory arthritis, psoriatic arthritis and spondylarthritis. The mean age of participants was 49.8 years (range 19 to 86 years), and most participants were female (57.4%). As presented in Table 1, 61% of the patients were rheumatoid factor and/or CCP positive. Mean ±SD symptom duration was 6.3 ± 3.1 months and mean ±SD time since diagnosis was 1.2 ± 2.3 months.

**TABLE 1.** Demographic and pain data.

|  |  |
| --- | --- |
| **Demographic** |  |
| Age (Mean, SD, Range) | 49.8 ±15.4 (19-86) |
| Gender F (n, %) | 35 (57%) |
| Ethnicity (n, %) |  |
| White British | 33 (54%) |
| Black or Black British | 11 (18%) |
| Asian or Asian British | 7 (12%) |
| Other | 10 (16%) |
| Smoker -Current or prev (n, %) | 23 (38%) |
| Unemployed (n, %) | 9 (15%) |
| BMI (Mean, SD) | 26.8 (5.7) |
| **Diagnosis** |  |
| EIA (n, %) | 16 (26%) |
| RA (n, %) | 38 (62%) |
| PsA (n, %) | 6 (10%) |
| SpA (n, %) | 1 (2%) |
| Seropositive -RhF or CCP (n, %) | 37 (61%) |
| Time since diagnosis, months (Mean, SD) | 1.2 (2.3) |
| Symptom duration, months (Mean, SD) | 6.3 (3.1) |
| **Steroid and DMARDs** |  |
| IM depomedrone in last 3 months (n, %) | 25 (41%) |
| PO prednisolone in last 2 weeks (n, %) | 13 (21%) |
| IV methylprednisolone | 2 (3%) |
| Two forms of steroids | 3 (5%) |
| No DMARDs (n, %) | 25 (41%) |
| Mono-therapy (n, %) | 33 (54%) |
| Combination therapy (n, %) | 3 (5%) |
| **Quality of life** |  |
| MSK HQ, Mean ±SD | 29 (11) |
| EQ5D, Mean ± SD | 0.58 (0.28) |
| EQ5D Total Health VAS, Mean ± SD | 59 (22) |
| RAID, Mean ± SD | 4.7 (2.6) |
| **Mental Health** |  |
| GAD 7, Mean ±SD | 7.4 (6.0) |
| Anxiety total, Score ≥5 (n, %) | 38 (62%) |
| Mild Anxiety, Score 5-9 (n, %) | 21 (34%) |
| Moderate Anxiety, Score 10-14 (n, %) | 7 (11%) |
| Severe Anxiety, Score ≥15(n, %) | 10 (16%) |
| PHQ9, Mean ±SD | 8.1 (6.4) |
| Depression total, Score $\geq 5$ (n, %) | 40 (66%) |
| Mild depression, Score 5-9 (n, %) | 17 (28%) |
| Moderate depression, Score 10-14 (n, %) | 13 (21%) |
| Moderately Severe depression, Score 15-19 (n, %) | 7 (11%) |
| Severe depression, Score $\geq 20$ (n, %) | 3 (5%) |
| PHQ15, Mean ±SD | 9.0 (5.5) |
| Somatic symptoms total, Score $\geq 5$ (n, %) | 49 (80%) |
| Mild somatic symptoms, Score 5-9 (n, %) | 25 (40%) |
| Moderate somatic symptoms, Score 10-14 (n, %) | 15 (24%) |
| Severe somatic symptoms, Score $\geq 15$ (n, %) | 9 (15%) |
| **Pain** |  |
| Total body pain, Mean ±SD | 5.5 (2.1) |
| Target joint pain, Mean ±SD | 5.1 (2.5) |
| **Widespread pain** |  |
| Widespread pain Index (WPI) score, Mean ± SD | 5.6 (2.9) |
| Widespread Pain Index (WPI) score of $\geq 7$, (n, %) | 19 (31%) |
| Fibromyalgia, (n, %) | 13 (21%) |
| Total pain > Target joint pain, (n, %) | 23 (38%) |
| T-SJC > 7, (n, %) | 12 (20%) |
| **painDETECT** |  |
| painDETECT, Mean ± SD | 12.7 (8.0) |
| Likely neuropathic pain (score of $\geq 19$), (n, %) | 15 (25%) |
| Possible neuropathic pain (score between 13-18), (n, %) | 9 (15%) |
| Unlikely neuropathic pain (score $\leq 12$), (n, %) | 37 (61%) |

Table showing patient demographics, diagnosis and medication alongside results of questionnaire data collected for quality of life, mental health, fibromyalgia criteria and painDETECT. (n=61). Abbreviations: BMI (Body mass index), EIA (Early inflammatory arthritis), RA (Rheumatoid arthritis), PsA (Psoriatic arthritis), SpA (Spondylarthritis), RhF (Rheumatoid factor), CCP (cyclic citrullinated peptide), IM (Intra-muscular), PO (Oral), IV (Intra-venous), DMARDS (Disease modifying anti-rheumatic drugs), MSK HQ (Musculoskeletal health questionnaire), EQ5D (Euro-Quality of Life 5Dimensions), RAID (Rheumatoid arthritis impact of disease questionnaire), GAD 7 (Generalised anxiety disorder 7 questionnaire), PHQ9 (Patient health questionnaire 9), PHQ15 (Patient health questionnaire 15).

#### Disease activity and therapy

The mean ± SD DAS28-CRP was 3.8 ± 1, consistent with low to moderate disease activity (30), with mean 28 tender joint count (TJC) 6.5±5.2 and swollen joint count (SJC) 2.7±2.5. Total and target joint mean ±SD pain on an NRS scale (0–10) was 5.5 ±2.1 and 5.1±2.5 respectively.

37 (61%) had received steroids in some form at the time of assessment and 36 (59%) received cDMARDs, with methotrexate being the most frequently prescribed (n=32, 52%) (Table 1).

#### Mental health and quality of life

The prevalence of participants with patient-reported outcome indications of anxiety, depression, and somatisation in the patient population was high, with 38 individuals (62%) experiencing anxiety, 40 individuals (66%) experiencing depression and 49 individuals (80%) experiencing somatic symptoms (Table 1). Approximately 15% had severe levels of anxiety, somatisation or moderately severe/severe depression (Table 1 & Figure 1).

**FIGURE 1:**
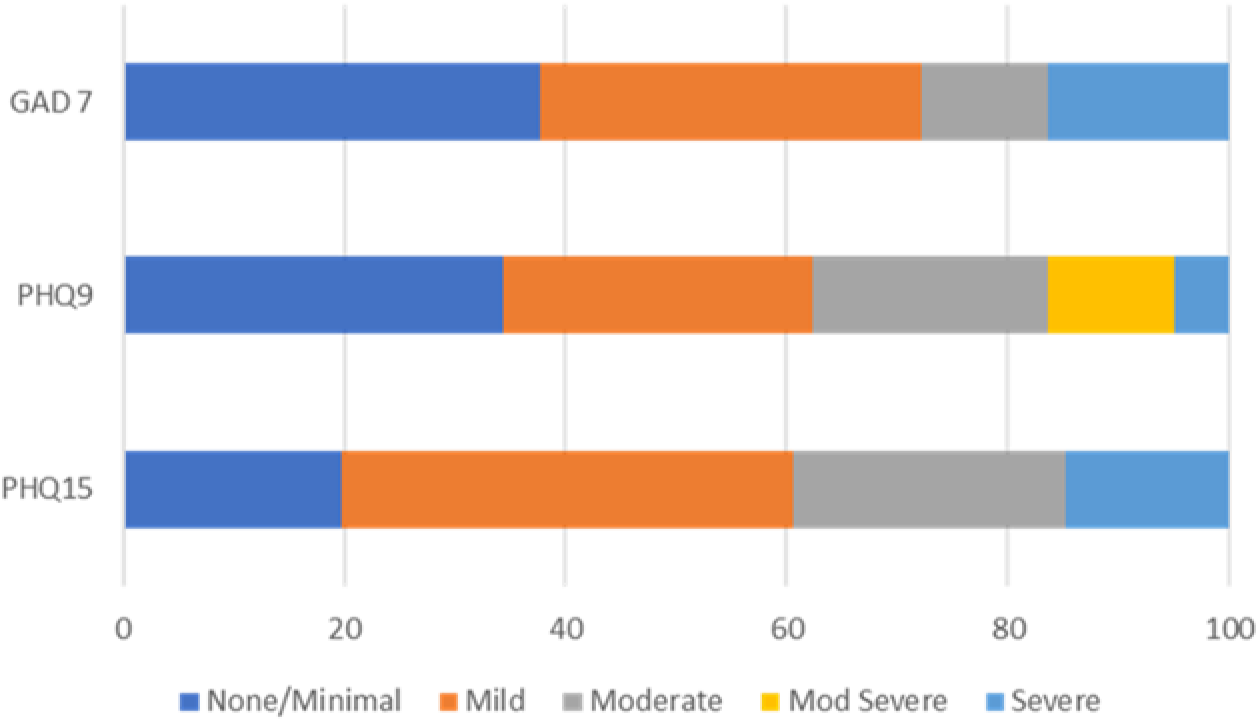
Prevalence of anxiety (GAD 7), depression (PHQ9), and somatic symptoms (PHQ 15) in the study population (n=61). Abbreviations: GAD 7 (Generalised anxiety disorder 7 questionnaire), PHQ9 (Patient health questionnaire 9), PHQ15 (Patient health questionnaire 15).

Quality of life (Qol), assessed using MSK-HQ and EQ5D, and impact of disease, assessed using RAID (Table 1), was in keeping with established RA populations (31).

#### Ultrasonographic (US) assessment

There was evidence of active synovitis either clinically or on US in 59 (97%) patients in keeping with a recent IA diagnosis. 37 (61%) had GS positivity and 22 (36%) had PD positivity at the target joint (Table 2). Of the 24 (39%) patients who had neither GS or PD positivity at the target joint, 17 had GS or PD positivity at other joints and 7 did not, though 5 of those had clinically swollen joints not examined by US. Of note, 24 of the 39 patients with PD 0 at the target joint had recently received steroid therapy, which may have supressed the PD signal.

**TABLE 2:** Grey Scale and Power Doppler activity on musculoskeletal ultrasound of the target most painful joint. N (%) of patients with each score at the target joint.

| **Grey scale score** | **N (%)** | **Power doppler score** | **N (%)** |
| --- | --- | --- | --- |
| GS 0 | 24 (39%) | PD 0 | 39 (64%) |
| GS 1 | 16 (26%) | PD 1 | 5 (8%) |
| GS 2 | 18 (30%) | PD 2 | 11 (18%) |
| GS 3 | 3 (5%) | PD 3 | 6 (10%) |

#### Clinical and patient-reported outcome measures (PROMs)

The assessment of widespread pain index (WPI), Tender minus Swollen joint count and total pain provide an indication of widespread pain (34), which is often associated with centrally mediated pain. 19 patients (31%) had a WPI score of ≥7, and 12 (19.6%) exhibited a T-SJC score of ≥7, consistent with a widespread pain phenotype (32). Additionally, 23 (38%) patients reported experiencing more total body pain compared to pain specifically in the target joint (assessed using the NRS 0-10 rating) (Table 1).

13 (21.3%) patients met the diagnostic criteria for fibromyalgia, a prototypical ‘central sensitivity’ syndrome, and 15 (24.6%) patients had a score of ≥19, on painDETECT questionnaire, indicative of neuropathic-like pain.

### Quantitative sensory testing

Below is a summary of the QST findings. Further details for interested readers are presented in supplementary material.

#### Static QST: Cutaneous sensation and pain thresholds

There was no difference in cutaneous sensation (MDT) using standardised von Frey hairs or pain thresholds (MPT) using a standardized pinprick, compared to normative data at either the target joint (test) or control site (Figure 2A, Z Values <1.96). There was a significant increase in sensitivity in MPT at the control site compared to the target joint (p=0.006), which may reflect a normal difference in sensation between the joint and forearm sites, or hypoesthesia due to inflammation at the joint. Comparison with healthy controls would be required to determine this.

**Figure 2.**
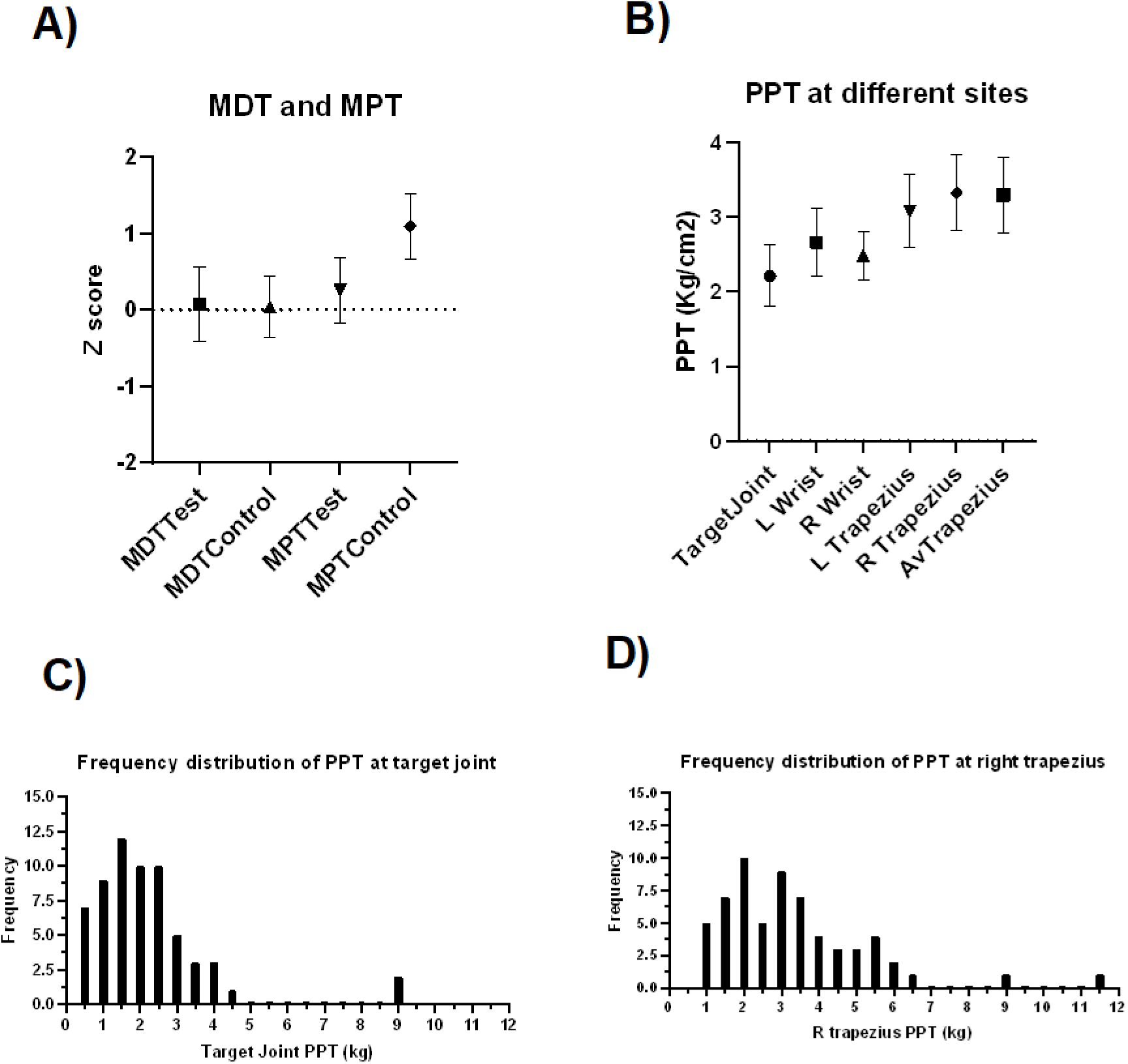
Graphs showing results from static quantitative sensory testing A) Z score for the study population mechanical detection threshold (MDT) and mechanical pain threshold (MPT) at test (target joint) and control sites (contra-lateral forearm) n=61. The QST z score chart illustrates the trend of sensory change where positive z scores suggest increased sensory capability (i.e., hypersensitivity), while negative scores point to diminished sensory ability (i.e., hyposensitivity). There is no significant difference in the MDT or MPT compared to normative data, reflected by Z scores < 1.96. B) Mean ± 95% confidence intervals for the PPT in (kg/cm^2^) at target joint, bilateral wrist and bilateral trapezius sites. n=60 C) Histogram of frequency distribution of PPT (kg/cm^2^) at the target joint n=60 D) Histogram of frequency distribution of PPT (kg/cm^2^) at the trapezius n=60

#### Static QST: Pressure pain thresholds (PPT) at target joint and trapezius

Low PPT reflects pain sensitisation. PPT values at the target joint were right skewed, with mean PPT at the target joint significantly lower than at the trapezius (p < 0.05), indicating peripheral sensitisation at the joint (Figure 2B, C). There was also right skewness in PPT at the trapezius, reflective of widespread hyperalgesia (Skewness at target joint 2.6, skewness at right trapezius 1.7) (Figure 2D).

#### Dynamic QST: Temporal summation of pain

Using the cuff algometer, 9/51 (18%) fulfilled criteria for facilitated TSP, with a ratio of >2.48 (Figure 3A). In contrast, there was no difference in TSP using pinpricks, compared to normative data at either the target joint (test) or control site. (Figure 3B).

**Figure 3.**
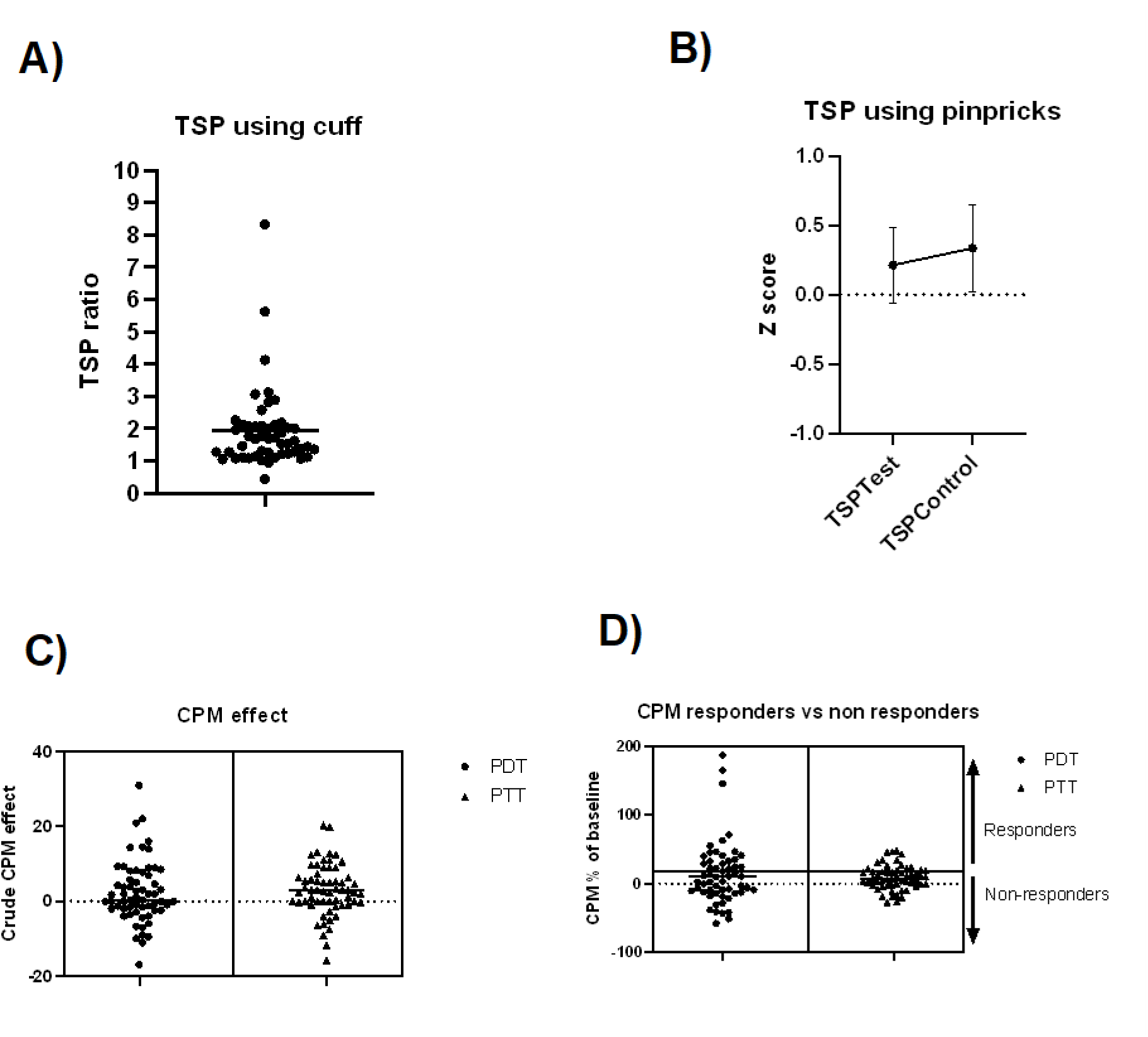
Figure 3 Graphs showing results from dynamic quantitative sensory testing A) Individual values for the temporal summation of pain (TSP) using cuff n=51. 9/51 (18%) fulfilled the criteria for facilitated TSP, with a ratio of >2.48. B) Z score for the study population temporal summation of pain (TSP), using pinprick n=51. There was no significant difference in TSP using pinpricks compared to normative data, reflected by Z scores < 1.96. C) Graph showing individual values of crude CPM effect (=PDT/PTT_(CPM)_- PDT/PTT _(Dom)_) for pressure detection threshold (PDT) and pressure tolerance threshold (PTT) n=57. D) Graph showing individual values of CPM effect as a % of baseline for PDT and PTT (Participants were stratified as responders (CPM response > 20% of the baseline cPPT) or non-responders (CPM response ≤20% of baseline cPPT) n=57. 35/57 (61%) according to PDT and 44/57 (77%) according to PTT were non responders.

#### Dynamic QST- Conditioned pain modulation

CPM non responders (i.e., those not reporting a decrease in pain perception upon application of a painful test stimulus concurrent to a conditioning stimulus) were recorded for pain detection threshold (35/57, 61%) and pressure tolerance thresholds (44/57, 77%) (Figure 3C, D). While pressure detection thresholds did not vary across CPM non responder and responder groups, CPM non-responders reported lower pressure tolerance thresholds, highlighting that although both groups had similar sensory detection, it was altered pain detection which stratified individuals into the non-responder group.

### Correlations and Factor analysis of clinical and patient reported outcome measures

Clinical and patient reported outcomes that suggest centrally mediated pain and lower non-articular PPT correlated with higher disease activity (DAS28), increased fatigue, decreased QoL, greater impact of disease and worse mental health measures (Table 3). In contrast, clinical measures of peripheral inflammation (total GS and PD positivity on US, swollen joint count), which suggest peripheral pain did not (Table 3).

**TABLE 3:** Correlation between markers of central pain/peripheral inflammation and clinical parameters. *p<0.05.

|  | DAS 28 | TJC | SJC | Time since diagnosis | Symptom duration | Fatigue | MSK-HQ | RAID | GAD-7 | PHQ-9 | PHQ-15 |
| --- | --- | --- | --- | --- | --- | --- | --- | --- | --- | --- | --- |
| **Markers central pain** |  |  |  |  |  |  |  |  |  |  |  |
| Total pain | 0.513 | 0.394 | 0.006 | -0.103 | 0.116 | 0.460 | -0.475 | 0.580 | 0.301 | 0.307 | 0.386 |
| Fibromyalgia severity (WPI+SSS) | 0.532 | 0.524 | -0.093 | -0.153 | 0.039 | 0.647 | -0.653 | 0.684 | 0.636 | 0.685 | 0.674 |
| PainDETECT | 0.514 | 0.454 | -0.132 | -0.175 | -0.073 | 0.501 | -0.500 | 0.596 | 0.501 | 0.493 | 0.681 |
| PPT at the trapezius (average) | -0.293 | -0.267 | 0.082 | 0.043 | 0.098 | -0.301 | 0.333 | -0.432 | -0.188 | -0.295 | -0.286 |
| **Markers of peripheral inflammation** |  |  |  |  |  |  |  |  |  |  |  |
| SJC | 0.337 | 0.207 | NA | 0.038 | 0.077 | 0.022 | -0.112 | 0.032 | -0.243 | -0.191 | -0.125 |
| Total GS & PD positivity | -0.106 | -0.007 | 0.546 | -0.019 | -0.001 | -0.021 | -0.036 | -0.039 | -0.210 | -0.222 | -0.199 |

Higher total pain, fibromyalgia severity, painDETECT scores and lowered PPT at the bilateral trapezius (taken as an average of both sides) correlated with higher disease activity (DAS28) and tender joint count (TJC), but not swollen joint count (SJC). They also correlated with increased fatigue, decreased QoL (MSK-HQ) and greater impact of disease (RAID). Additionally, these measures showed correlations with mental health measures, including higher levels of anxiety (GAD-7), depression (PHQ-9) and somatic symptoms (PHQ-15) (with the exception of PPT at the trapezius and anxiety). Notably, these measures did not demonstrate correlations with either time since diagnosis or symptom duration. Total GS and PD positivity in the 14 joint ultrasound assessment represents the combined scores of grey-scale and power doppler assessments. The swollen joint count (SJC) and total GS and PD positivity serve as indicators of peripheral inflammation, which contributes to peripheral pain. SJC correlated with disease activity (DAS28) and total GS and PD positivity correlated with SJC. However, neither the SJC or total GS and PD positivity correlated with TJC, fibromyalgia severity, QoL by MSK-HQ or RAID or mental health measures (GAD-7, PHQ-9 or PHQ-15). Those highlighted in green are statistically significant at the $p < 0.05$ level, whereas those highlighted in orange are not.

Based on factor analyses of the 14 variables, we identified two underlying latent factors. CRP, TSP and CPM had KMO statistics below 0.5 and were not included in the model. The overall KMO statistic was 0.76 indicating the correlation matrix of the remaining variables was appropriate for factor analysis. Factor 1 included self-reported variables of pain (total pain, patient global assessment, TJC, painDETECT, fibromyalgia criteria and trapezius PPT), fatigue and mental health (PHQ 9 and GAD 7). Factor 2 included clinical and ultrasound measures of synovitis (SJC, GS and PD on MSK US and physician global assessment) (Table 5, Supplementary material). The correlation between the two factors was -0.13.

## DISCUSSION

This study provides novel insights into the mechanisms underlying pain reported by patients with early IA, as we compiled comprehensive sensory profiles of individuals shortly after diagnosis. Consistent with newly diagnosed IA, 97% of patients had active synovitis confirming that peripheral inflammation is a major contributor to pain at this stage. However, we also observed indicators of centrally mediated pain in approximately 20-30% of patients, who reported widespread pain, fulfilled diagnostic criteria for fibromyalgia, and/or demonstrated elevated painDETECT scores. QST findings further supported the involvement of centrally mediated pain mechanisms. Reduced pressure pain thresholds (PPT) at non-articular sites in some patients, suggested widespread hyperalgesia, while heightened responses to temporal summation of pain (TSP) and impaired conditioned pain modulation (CPM) indicated dysfunction in spinal and supraspinal processes. Overall, these data challenge our current understanding that centrally mediated pain processes are absent at diagnosis and develop over disease course. Instead, they suggest that some patients with early IA have both inflammatory and centrally mediated pain, similar to patients with established IA (5,13,33)

Our cohort’s clinical disease activity measures and average pain score of 5.5/10 aligns with findings reported in other early inflammatory arthritis cohorts (34), as do the comparably high rates of anxiety and/or depression (35) (36). While pain mechanisms in early IA remain relatively unexplored, our findings revealed that 21% of patients met the criteria for fibromyalgia—far exceeding the 5.2% reported in one Canadian study (37). This may reflect methodological differences, with the Canadian study relying on clinical judgment rather than strict fibromyalgia diagnostic criteria (37) or that the Canadian cohort had a more inflammatory profile (36) than ours or other early IA cohorts (37,38). In addition, symptoms of very active early IA, such as widespread pain, fatigue and unrefreshing sleep, overlaps with fibromyalgia criteria, potentially resulting in an “false” fibromyalgia diagnosis. However, the lack of correlation between WPI/fibromyalgia severity and peripheral inflammation markers suggests that the observed widespread pain may be driven by mechanisms other than joint inflammation.

We observed reduced PPT values at the affected joint in most patients indicating local pain sensitisation, likely due to joint inflammation, in this early arthritis cohort. In addition, we found lowered PPT at non-articular sites, indicative of widespread pain, in some patients. These findings are in keeping with those from a recent meta-analysis of patients with established IA (13), but are the first to identify such reductions in PPT at non-articular sites specifically in the context of early IA, highlighting the presence of widespread pain sensitivity in some patients early in the disease course. Factor analysis revealed close links between reduced PPT at non-articular sites with self-reported pain, fatigue and poor mental health which are often associated with centrally mediated pain. The lack of interaction with dynamic QST (TSP or CPM) measures, suggests that reduced PPT at non articular sites may be the most robust QST indication of centrally mediated pain in this patient population.

18% of participants demonstrated facilitated TSP, when a TSP paradigm with the cuff algometer was used as a repetitive stimulus, indicating spinal mechanisms contributing to heightened pain. However, when pinpricks were used as the repetitive stimulus, facilitated TSP was not observed; contrasting with findings from a study that also used pinpricks in patients with established IA (39). This difference in findings could be attributed to characteristics specific to our study population – ours was a recently diagnosed IA patient cohort - or alternatively, due to methodological protocols. For example, the initial pinprick force may not have been sufficiently noxious, or the initial pinprick rating may have been too high, creating a ceiling effect on subsequent pain ratings. When applying a CPM paradigm, 61% and 77% of the participants exhibited a ‘non-responder status’ according to pressure detection threshold and pressure tolerance threshold respectively, indicating dysfunction in a naturally occurring pain inhibitory pathway. Although data is lacking to compare the proportion of non-responders with healthy controls (40), our cohort’s mean CPM effect for PDT was 2.4 (supplementary material), below that found in healthy populations where the mean CPM effect for PDT ranges from 5.3-11.5 (27)

This study has several strengths. The in-depth sensory profiling allowed us to create a comprehensive profile, combining measures of pain sensitivity and centrally mediated pain processing mechanisms, and ultrasound as an objective measure of joint inflammation. All assessments were carried out by a single researcher, ensuring consistency and minimising inter-rater variability. We attempted early assessment of patients soon after their diagnosis, which was usually achieved, enabling insights into what is driving pain in the very early stages of IA soon after diagnosis.

The study also has limitations. One key limitation is that there is currently no “gold standard” comprehensive method for assessing centrally mediated pain. As a result, instruments such as questionnaires and QST, which have been validated individually, are combined. However, these instruments are frequently not standardised or validated in comparison to each other. For example, although used in the literature as a proxy measure of centrally mediated pain, there is currently limited data linking “likely neuropathic pain” identified by painDETECT to QST findings in IA [45,46]. At the time of study assessment most patients had already received therapy and while we captured patients within a short time post-diagnosis, most individuals had pain in the period prior to diagnosis, and so in some individuals, central nervous system dysfunction may have pre-dated joint inflammation. It has been postulated that a pre-loading psychological vulnerability to pain may contribute to the development of persistent non-inflammatory pain. An alternative explanation is that this could also be influenced by sub-clinical joint pathology. We will follow this group with longitudinal investigations aimed at identifying evolution of pain mechanisms over time and possible signals for those who might develop persistent non-inflammatory pain. [46,47]

## Conclusion

These findings emphasise that pain in early IA is complex and, in some people, includes centrally mediated pain mechanisms, highlighting the need for awareness of these factors in early inflammatory arthritis populations. Future longitudinal studies are essential to track the evolution of pain mechanisms over time and identify predictors of persistent pain, which may inform early interventions targeting central pain processes.

## Data Availability

All data is available upon request to the corresponding author.

## Acknowledgements

We thank Professor Thomas Graven Nielson for his invaluable advice and support in training and interpretation of cuff algometer data throughout the project.

## Data access statement

All data is available upon request to the corresponding author.

